# Paclitaxel Drug-coated Balloon Angioplasty for De Novo Coronary Lesions in an Expanded Real World Clinical Setting: The Multicenter ALLIANCE Registry

**DOI:** 10.64898/2025.11.30.25341329

**Authors:** Masato Nakamura, Kengo Tanabe, Kazushige Kadota, Takashi Muramatsu, Yutaka Tadano, Kenji Ando, Shigeru Nakamura, Takashi Ashikaga, Yoshihisa Kinoshita, Nehiro Kuriyama, Yuko Onishi, Toru Kataoka, Koji Nishida, Raisuke Iijima, Masatsugu Nozoe, Kunio Morishige, Takefumi Takahashi, Yoshitaka Murakami, Ken Kozuma, the ALLAINCE registry investigators

**Affiliations:** Division of Minimally Invasive Treatment in Cardiovascular Medicine, Toho University Ohashi Medical Center, Tokyo, Japan; Division of Cardiology, Mitsui Memorial Hospital, Tokyo, Japan; Department of Cardiovascular Medicine, Kurashiki Central Hospital, Kurashiki, Japan; Department of Cardiology, Fujita Health University Hospital, Toyoake, Japan; Department of Cardiology, Sapporo Cardio Vascular Clinic, Sapporo, Japan; Department of Cardiology, Kokura Memorial Hospital, Kitakyushu, Japan; Cardiovascular Center, Kyoto Katsura Hospital, Kyoto, Japan; Department of Cardiology, Japanese Red Cross Musashino Hospital, Tokyo, Japan; Department of Cardiovascular Medicine, Toyohashi Heart Center, Toyohashi, Japan; Department of Cardiology, Miyazaki Medical Association Hospital, Miyazaki, Japan; Department of Cardiology, Hiratsuka Kyosai Hospital, Hiratsuka, Japan; Division of Cardiology, Bell Land General Hospital, Sakai, Japan; Department of Cardiology, Chikamori Hospital, Kochi, Japan; Division of Cardiovascular Medicine, Toho University, Ohashi Medical Center, Tokyo, Japan; Division of Cardiology, Saiseikai Fukuoka General Hospital, Fukuoka, Japan; Department of Cardiovascular Medicine, Matsuyama Red Cross Hospital, Matsuyama, Japan; Department of Cardiovascular Medicine, Tokushima Red Cross Hospital, Komatsushima, Japan; Department of Preventive Medicine and Public Health, Toho University School of Medicine, Tokyo, Japan; Division of Cardiology, Teikyo University Hospital, Tokyo, Japan

**Keywords:** drug-coated balloon, real world, percutaneous coronary intervention, imaging guide

## Abstract

**Background:** Despite limited clinical evidence, the use of drug-coated balloons (DCB) in percutaneous coronary intervention for de novo coronary lesions is expanding and increasing. Furthermore, recent randomized trial demonstrated that imaging guided DCB improves angiographic outcomes.

**Methods:** ALLIANCE is a prospective, multicenter, all-comer registry designed to evaluate the efficacy and safety of the paclitaxel-DCB in an expanded real-world clinical setting. A calculated sample size of 1500 patients was expected to provide 95% power to detect non-inferiority against a performance goal of 7.5% target lesion failure (TLF) at 1 year with a 3.0% absolute non-inferiority margin. Cases in which DCB was performed after successful lesion preparation were consecutively enrolled. The Primary endpoint was TLF, a composite of cardiac death, myocardial infarction and clinically driven target lesion revascularization (cd-TLR) at 1 year. An independent clinical events committee adjudicated all primary endpoint events.

**Results:** Between July 2023 and February 2024, 1794 patients (with 1984 lesions) were enrolled at 57 sites in Japan. Mean age was 71 years, 76% of patients were male, 45% had diabetes mellitus, and 29% presented with acute coronary syndromes. Among lesion characteristics, 31% were calcified lesions, 25% showed diffuse disease, 26% were bifurcations, and 38% were treated a greater than 3 mm DCB balloon. Intravascular imaging was used in 88.5% (1755/1984) of lesions. Lesion preparation was performed with modified balloons in 68% of cases and by atherectomy in 19%. Bail-out stenting after DCB was required in 1.1% of patients. The rate of TLF at 1 year was 4.7% overall (95%CI 3.8-5.8%, p_non-inferiority_<0.001), and cd-TLR 2.9%. Thrombotic events during the follow-up period were limited to one case

**Conclusions:** Paclitaxel DCB has proven effective and safe in the current expanding daily clinical practice. Utilizing imaging modalities during DCB procedure may improve DCB treatment outcomes.

**What is Known?:**

- Clinical applications of paclitaxel DCBs are being explored for a variety of lesion subsets and clinical presentations; however, their expanded clinical use has not been validated in large-scale prospective studies.
- Guidance from imaging devices may enhance the potential of DCBs in routine clinical practice; however, no data demonstrate clinical benefit from the combined use of DCBs and imaging modality.

**What the study adds?:**

- Paclitaxel DCB was effective and safe in various coronary artery lesion and clinical settings in real world practice.
- The use of an imaging modality may enable optimal preparation of lesions, increasing the inherent potential of DCB in daily practice.

## Introduction

The advent of drug-eluting stents (DES) has seen a significant reduction in restenosis rates, and percutaneous coronary intervention has now assumed a central role in the treatment of ischemic heart disease. Nevertheless, because DES permanently cage vessels, they have significant limitations in terms of long-term outcomes.^1,2)^ Therefore, drug coated balloons (DCBs) have been investigated numerous clinical trials,^3,4)^ and recent reports indicate DCBs are effective in acute coronary syndrome (ACS) and non-small vessels.^5,6)^ Therefore, it is presumed that the use of DCB is continuing to expand at present. Nevertheless, large real-world studies of broad clinical use remain limited and further evaluation is necessary. Furthermore, the ULTIMATE III trial recently suggested the clinical utility of imaging guided DCB procedures over angiography guided DCB from the perspective of angiographic parameters.^7)^ However, the clinical benefits of the use of imaging modalities at the time of DCB procedures in real-world practice remain unclear. Here, we evaluated the efficacy and safety of paclitaxel DCB therapy combined with an imaging modality in the currently broad clinical setting and lesion types.

## Methods

### Data Availability Statement

Data will not be available to others.

### Study population

The ALLIANCE registry is an all-comer prospective multicenter registry which is examining the efficacy of DCB for today’s expanded indications. Inclusion criteria were age 18 years or older, written consent to participate in this study, and consideration as suitable for DCB treatment. Exclusion criteria were limited to participation in other clinical trials, consideration as unsuitable for DCB by the treating physician, and in-stent restenosis. Cases treated with DCB were consecutively enrolled, however, cases where a physician determined that lesion preparation had failed were treated with DES implantation without the use of DCB and were not enrolled. The study protocol was approved by central review (Ethical Committee of Toho University Ethics Board at Toho University (A23029, 7 July 2023), and the study was registered in Japan Registry of Clinical Trials (jRCT1032230218).

### DCB procedure with the use of an imaging modality

All DCB procedures were performed according to current guidelines and local practices. ^8)^ Regarding lesion preparation, residual stenosis visually less than 30% and absence of major dissection compromising blood flow after pre-dilatation was defined as adequate angiographic findings. ^8)^ Imaging modalities were used to select the preparation device based on plaque characteristics, determine the pre-dilatation balloon size, establish the lesion length to be covered with the DCB, and confirm acute lumen gain after lesion preparation. A balloon size corresponding to a 1:1 ratio based on the lumen diameter in areas with a plaque burden less than 50% is generally recommended.^8)^ However, since no consensus has been reported regarding optimal lesion preparation criteria based on imaging examinations,^9)^ the determination of optimal lesion preparation was left to the discretion of each physician. In cases where coronary artery flow-limiting dissection occurred during dilation with a DCB and dilation was inadequate, bail-out stenting was recommended without hesitation. Only two types of DCB were available in Japan, both paclitaxel-coated balloons, namely SeQuent® Please NEO (B. Braun, Melsungen, Germany) and AGENT^TM^ (Boston Scientific, MA, USA). The choice of DCB was left to the operator. Multiple overlaps were allowed. DCBs with a length of 2 mm covering both ends were selected to avoid lesion mismatch. Dilatation was routinely set for at least 1 min. A hybrid strategy, defined as overlapping DES and DCB for diffuse coronary artery lesions or bifurcation lesions, was allowed.

### Follow-up

Dual antiplatelet treatment (DAPT) was basically administered according to the device’s instructions for use, but the duration of DAPT was at the discretion of the physician. Clinical follow-up was scheduled at 6, 12, 24 and 36 months, with the 6- and 12-month follow-ups conducted at the institution while the 24- and 36-month follow-ups could be conducted by telephone and letter. Coronary angiographic follow-up was not mandatory but rather based on clinical need essentially at the discretion of the individual center.

### Endpoints

The primary endpoint of the study was the incidence of target lesion failure (TLF) at 1 year (365 ± 30 days) post-procedure, defined as a composite of clinically driven target lesion revascularization (cd TLR) based on clinical findings, target vessel-related myocardial infarction or cardiac death. Cd TLR was defined as revascularization based on symptomatic or functional stenosis assessment or positive ischemia on stress testing, or stenosis of ≥70% on quantitative coronary angiography. Technical success was defined as post procedural residual lesion stenosis of <50% without severe coronary dissection, and clinical procedural success was defined as meeting the criteria for technical success and no death or myocardial infarction within 24 hours of the procedure. Other endpoints included individual components of the TLF, all-cause mortality, target vessel failure (TVF) and bleeding events as defined by the Bleeding Academic Research Consortium (BARC).^10)^ Events associating with primary and key secondary endpoints were adjudicated by an independent clinical event committee. (Table S1) Details of the definitions are provided in the Table S2.

### Statistical analysis

This prospective registry was designed with sufficient sample size to assess the efficacy and safety of DCB in real world settings. Referencing previous reports, an assumed performance goal of 7.5% for TLF was set^4)^ and a clinically meaningful difference, considered to be below the non-inferiority margin, was 3%. We set 2.5% for the type 1 error and 95% power for our non-inferiority test, and calculated a sample size of 1,231 cases. We determined that the minimum number of patients should be 1,500, considering the expected attrition rate, and that the maximum should be 2,000, considering real-world practice. Categorical variables were expressed as a percentage with respect to relative frequency and analyzed using the Chi-square test or Mann-Whitney U test. Continuous variables were expressed as mean ± standard deviation or as median if not normally distributed and analyzed using the t-test or Fisher exact test, respectively. Clopper-Pearson 95% confidence intervals (CIs) were constructed. In this study, non-small vessels were defined by a DCB size ≥3 mm. Variables attributable to TLF and cd-TLR were examined using multivariate Cox regression analysis. Adjustment variables included those considered clinically important, such as age, gender, diabetes mellitus (DM), and dyslipidemia, plus those with a difference of p<0.1 on univariate analysis. Multicollinearity was assessed using variance inflation factors and evaluated using coefficients, all of which were less than 5. Consequently, all coefficients were less than 1.2. The choice of variables to be analyzed in the model followed statistical and then clinical criteria. p values of <0.05 from two-tailed testing were considered to indicate statistical significance. All analyses were performed using SAS Version 9.4 (SAS Institute).

## Results

### Subject population

A flow chart of the study is shown in Figure 1. A total of 1817 patients from 57 institutions were enrolled between July 2023 and February 2024 (Table S3). This accounted for 15% of all treated lesions during this period, excluding in-stent restenosis lesions, and 70.5% of all lesions treated by DCB. Among these patients, 23 patients were excluded from analysis, while the remaining 1794 patients were included in the study analysis. Follow-up rate at 1 year was 97.3%. Patient background is shown in Table 1. 76.3% of subjects were male and mean age was 70.5 years. By incidence, 44.5% of patients had DM, 28.6% had ACS, and 8.2% were receiving hemodialysis. More than half of all patients were categorized as at high bleeding risk by the Academic Research Consortium definition. ^11)^ Lesion background and procedural characteristics are shown in Table 2. The main target vessel for treatment was the left anterior descending artery (41.0%). Lesions in the left main trunk artery accounted for 1.6% and de-novo lesion for 97.4%. Ostial lesions, calcified lesions and diffuse lesions accounted for 21.0%, 31.2%, and 25.0%, respectively, while 25.6% of lesions were bifurcated.

**Fig 1.**
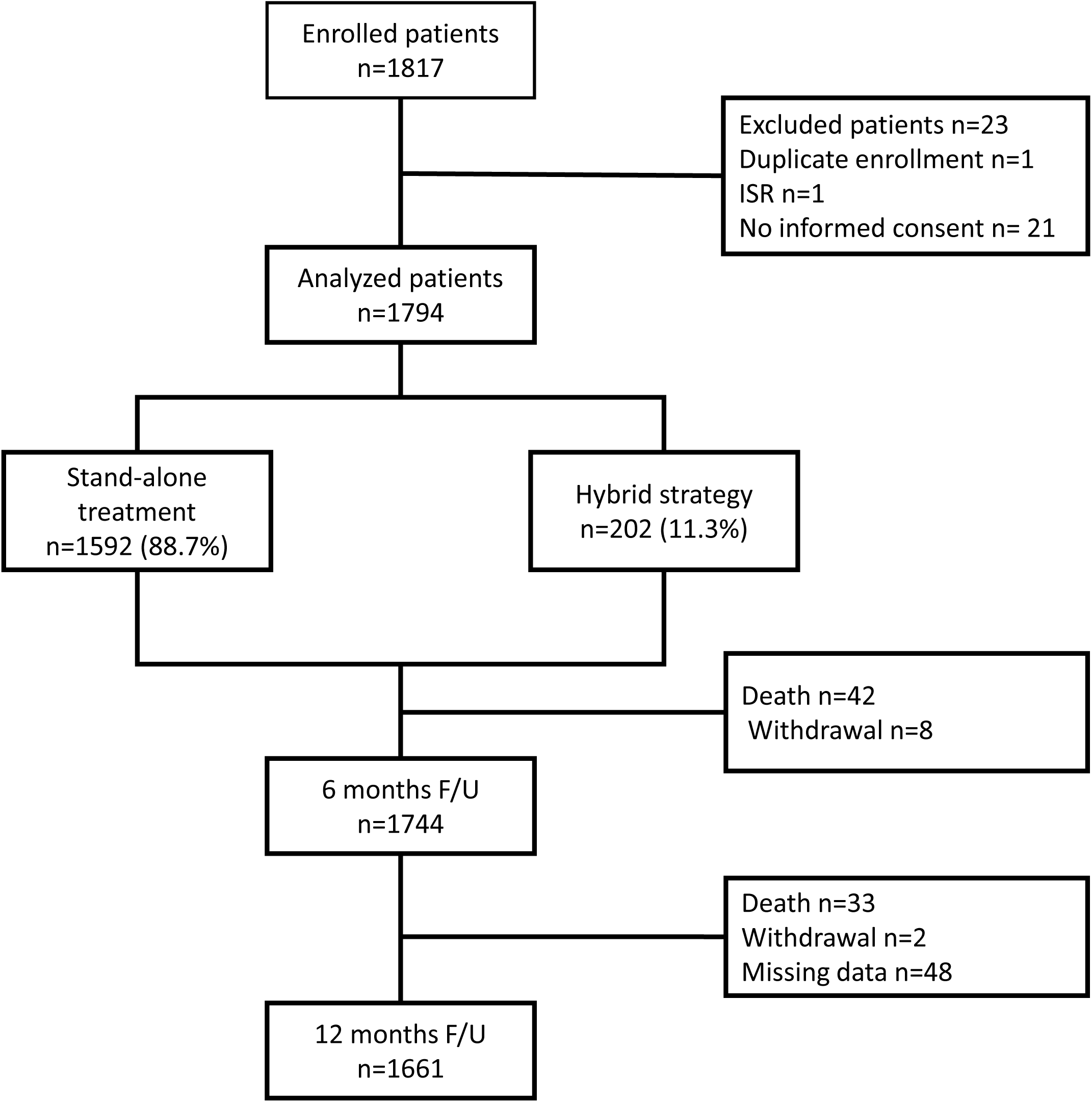
Flow chart of the study. A total of 1817 patients were enrolled between July 2023 and February 2024 from 57 institutes. 1794 patients were included in this study analysis and follow-up rate at 1year was 97.3%. ISR: in stent restenosis

**Table 1:** Patient demographics.

|  |  |
| --- | --- |
| No. of patients | 1794 (%) |
| **Demographics** |  |
| Age, years | 70.5±11.6 |
| ≥75 years | 764 (42.6) |
| Sex (Male) | 1369 (76.3) |
| Height (cm) | 162.9±9.3 |
| Wight (Kg) | 64.5±13.9 |
| Body mass index (kg/m<sup>2</sup>) | 24.1±4.0 |
| Smoking Current | 293 (16.3) |
| Former | 757 (42.2) |
| **Comorbid conditions** |  |
| Diabetes mellitus | 799 (44.5) |
| Insulin-treated diabetes | 145 (18.1) |
| Hyperlipidemia | 1395 (77.8) |
| Hypertension | 1388 (77.4) |
| Previous history of heart failure | 219 (12.2) |
| Atrial fibrillation | 188 (10.5) |
| On hemodialysis | 148 (8.2) |
| Liver cirrhosis | 10 (0.6) |
| Peripheral artery disease | 178 (9.9) |
| Previous history of PCI | 867 (48.3) |
| History of coronary artery bypass grafting | 65 (3.6) |
| Previous myocardial infarction | 444 (24.7) |
| Previous stroke | 173 (9.6) |
| Previous episode of blood transfusion | 21 (1.2) |
| Frailty score | 2.7±1.4 |
| High bleeding risk (ARC definition) | 1005 (56.0) |
| eGFR (ml/min/1.73m<sup>2</sup>) | 58.3±24.4 |
| Hb | 13.2±1.9 |
| Warfarin/DOAC | 32/163 (10.9) |
| **Clinical presentation** |  |
| Stable angina | 917 (51.1) |
| Silent ischemia | 364 (20.3) |
| Acute coronary syndrome | 513 (28.6) |
| ST-elevated myocardial infarction | 178 (9.9) |
| Non-ST-elevated myocardial infarction | 174 (9.7) |
| Unstable angina | 161 (9.0) |

Values are mean SD, n (%).
PCI, percutaneous coronary syndrome; ARC, Academic Research Consortium; DOAC, direct oral anticoagulant; eGFR, estimated glomerular filtration rate.

**Table 2.** Baseline lesion and procedural characteristics.

|  |  |
| --- | --- |
| No. of lesions | N=1984 |
| No. of treated lesions/patient | 1.1±0.3 |
| Lesion location |  |
| Left main trunk | 31 (1.6) |
| Right coronary artery | 517 (26.1) |
| Left anterior descending artery | 813 (41.0) |
| Left circumflex artery | 621 (31.3) |
| Bypass graft | 2 (0.1) |
| Lesion characteristics |  |
| De-novo lesion | 1932 (97.4) |
| Ostial lesion | 416 |
| Calcified lesion (>moderate) | 619 (31.2) |
| Thrombotic lesion | 171 (8.6) |
| Proximal vessel tortuous lesion | 158 (8.0) |
| Angulated lesion >90° | 127 (6.4) |
| Eccentric lesion | 680 (34.3) |
| Diffuse lesion (>20mm) | 496 (25.0) |
| Chronic total occlusion | 101 (5.1) |
| Bifurcated lesion | 507 (25.6) |
| Procedure |  |
| The use of an imaging modality | 1755 (88.5) |
| IVUS | 1524/1755 (86.8) |
| OCT | 231/1755 (13.2) |
| Type of pre-dilatation |  |
| Pre-dilatation | 1972 (99.4) |
| Plain old balloon | 885 (44.9) |
| Cutting balloon | 792 (40.2) |
| Scoring balloon | 546 (27.7) |
| Rotablator/orbital atherectomy | 183/81 (13.3) |
| ELCA | 32 (1.6) |
| Directional atherectomy | 84 (4.3) |
| Coronary dissection by pre-dilatation | 612 (31.0) |
| DCB |  |
| AGENT<sup>TM</sup> | 1445 (73.3) |
| SeQuent<sup>®</sup> Please NEO | 523 (26.4) |
| AGENT<sup>TM</sup>+SeQuent<sup>®</sup> Please NEO | 6 (0.3) |
| Dilatation time (sec) | 58.1±20.9 |
| Dilatation pressure (atm) | 7.0±2.2 |
| Coronary dissection after DCB | 267 (13.5) |
| Bail-out stenting | 22 (1.1) |
| Hybrid strategy | 202 (10.2) |

Values are mean SD, n (%).
IVUS, intravascular ultrasound; OCT, optical coherence tomography; DCB, drug coated balloon; ELCA, Excimer laser coronary angioplasty.

### Procedures

DCB with the use of imaging modalities was conducted in 88.9% of cases, mainly using intravascular ultrasound. A modified balloon, such as a cutting balloon and scoring balloon, was frequently used for lesion preparation (67.8%), while an atherectomy device was used in 19.2% of cases. Coronary dissection was observed in 31%, but the majority were National Heart, Lung, and Blood Institute type A or B, and severe coronary dissection exceeding type B^12)^ was limited to 67 lesions (3.4%) (Table S4).

Target lesions were treated by DCB with a mean balloon size of 2.7±0.6 (balloon/artery ratio 1.16±0.57) and length of 23.8 ± 10.8 mm. Bail out stenting was eventually required in 22 cases (1.1%). The use of antithrombotic drugs during the follow-up period is described in Table S-5. At discharge, 86.9% were prescribed DAPT and 11.9% were prescribed anticoagulants. Among patients prescribed DAPT, 41.9% received the agent for 6 months and 25.4% for 1 year.

### Primary and secondary endpoints

Clinical endpoint at 1 year is summarized in Table 3. TLF at 1 year was 4.7% (95% CI: 3.8–5.8). This rate falls below the non-inferiority margin assumed for the performance target TLF (p_non-inferiority_<0.001). The rates of cd TLR, myocardial infarction, and cardiac death, which comprise TLF, were 2.9%, 0.4%, and 1.7%, respectively. Acute coronary occlusion during follow-up was limited to 1 case. Major bleeding with BARC 3,5 was observed in 56 patients (3.1%) and clinically relevant bleeding with BARC 2,3,5 in 95 patients (5.3%).

**Table 3.** Clinical follow-up outcomes.

| Primary endpoint |  | 95% CI |
| --- | --- | --- |
| Target lesion failure | 85 (4.7) | 3.8-5.8 |
| Secondary endpoints |  |  |
| Any death | 75 (4.2) | 3.3-5.2 |
| Cardiac death | 30 (1.7) | 1.2-2.4 |
| Noncardiac death | 45 (2.5) | 1.9-3.3 |
| Any MI | 9 (0.5) | 0.2-1.0 |
| Target vessel MI | 7 (0.4) | 0.2-0.8 |
| Non-target vessel MI | 2 (0.1) | 0.0-0.4 |
| Any revascularization |  |  |
| Clinically driven TLR | 52 (2.9) | 2.2-3.8 |
| Clinically driven TVR | 60 (3.3) | 2.6-4.3 |
| Target vessel failure | 93 (5.2) | 4.3- 6.3 |
| Definite/probable vessel thrombosis | 1 (0.1) | 0.0 - 0.3 |
| All death, MI, TVR | 138 (7.7) | 6.5-9.0 |
| Cardiac death + MI | 38 (2.1) | 1.4-2.7 |

Values are mean SD, n (%). 95% confidence interval
MI, myocardial infarction; TLR, target lesion revascularization; TVR, target vessel revascularization.

### Determinants of TLF and cd-TLR

The results for 1-year TLF and cd-TLR in clinically important subgroups are provided in Figure 2 and Tables S 6 and S7. The results were relatively consistent and favorable in all subgroups. Cox regression multivariate analysis revealed that DM, hemodialysis and ostial lesion were independent risk factors for both TLF and cd-TLR. Clinical risk factors of ACS and hyperlipidemia was associated with TLF only. Of note, calcified lesion, non-small vessel and bifurcation were not associated with cd-TLR (Table 4, Table S6).

**Fig 2.**
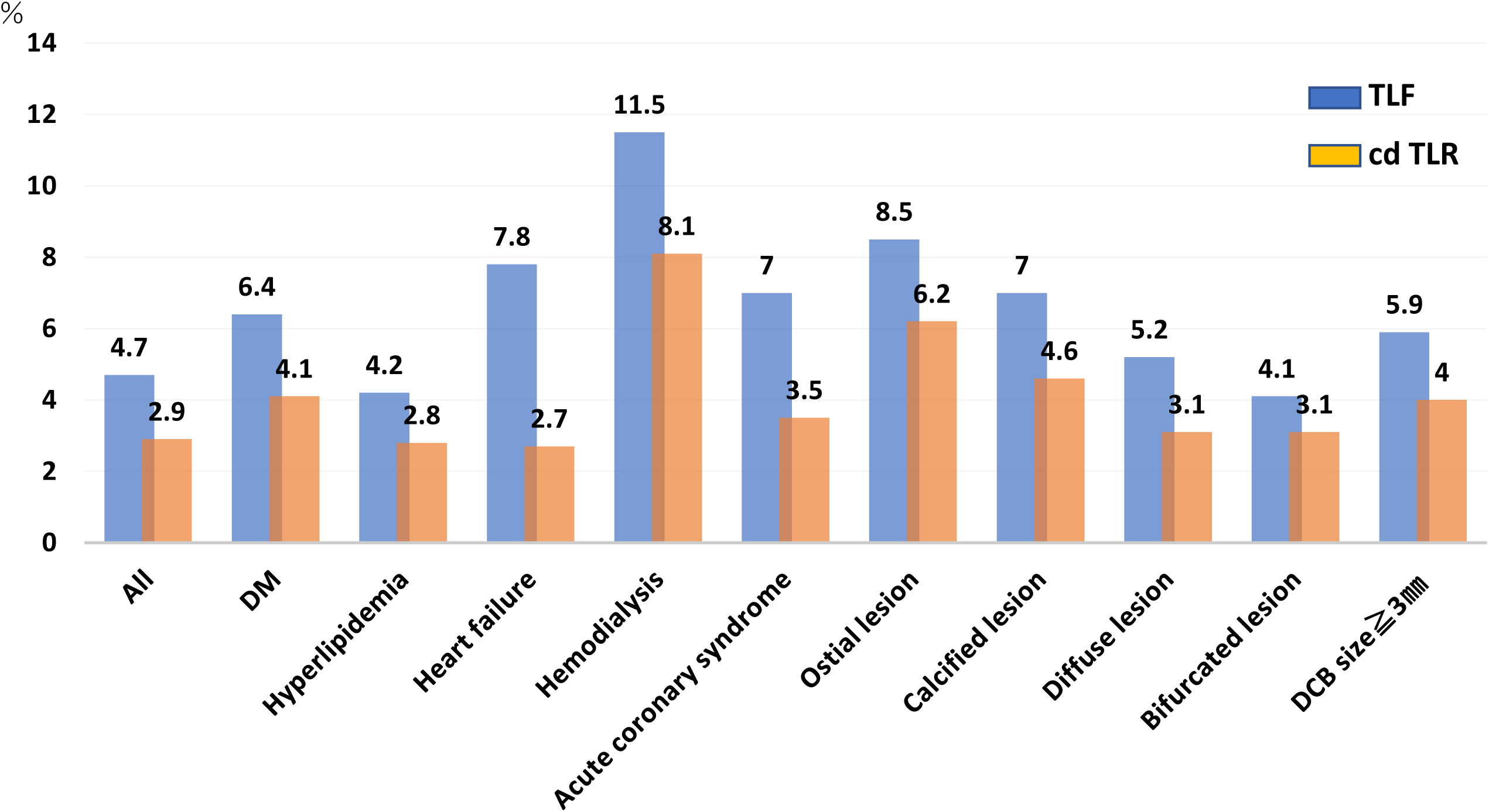
1-year TLF and TLR outcomes in various subgroups. TLF and cd-TLR were relatively consistent across various subgroups. DM: diabetes mellitus.

**Table 4:** Multivariate Cox regression analysis results of TLF and cd TLR.

|  | TLF |  |  | cd-TLR |  |  |
| --- | --- | --- | --- | --- | --- | --- |
|  | HR | 95%CI | p-value | HR | 95% CI | p-value |
| Age 65-74 yrs | 1.07 | 0.57-2.00 | 0.830 | 1.17 | 0.57-2.41 | 0.674 |
| Age $\geq 75$ yrs | 1.48 | 0.84-2.60 | 0.174 | 1.13 | 0.56-2.28 | 0.742 |
| Sex (Female) | 1.18 | 0.84-2.60 | 0.529 | 0.89 | 0.44-1.82 | 0.757 |
| Diabetes mellitus | 1.78 | 1.14-2.78 | 0.012 | 2.00 | 1.13-3.58 | 0.018 |
| Hyper lipidemia | 0.59 | 0.37-0.94 | 0.026 | 0.74 | 0.39-1.40 | 0.355 |
| Heart failure | 1.43 | 0.82-2.48 | 0.207 | 0.70 | 0.29-1.68 | 0.356 |
| Acute coronary syndrome | 2.06 | 1.33-3.18 | 0.001 | 1.54 | 0.87-2.75 | 0.142 |
| Hemodialysis | 2.25 | 1.26-4.01 | 0.006 | 2.58 | 1.27-5.27 | 0.009 |
| Ostial | 2.18 | 1.40-3.40 | <0.001 | 3.00 | 1.72-5.22 | <0.001. |
| Calcified | 1.37 | 0.86-2.19 | 0.184 | 1.58 | 0.86-2.89 | 0.139 |
| DCB size $\geq 3.0$mm | 1.44 | 0.93-2.23 | 0.099 | 1.56 | 0.90-2.72 | 0.115 |

cd TLR, clinically driven target lesion revascularization; HR, hazard ratio; TLF, target lesion failure.

## Discussion

This large-scale, multicenter DCB registry completed enrollment in just over 7 months and demonstrated that paclitaxel DCB yield favorable clinical outcomes in expanding clinical settings. Main findings are as follows: First, the 1-year TLF rate was low at 4.7% across various lesions and clinical scenarios. Second, only 1.1% of cases required bail out stenting. Third, DM, hemodialysis, and ostial lesions were independent risk factors for both TLF and cd-TLR.

Recent findings have suggested efficacy of DCB in a wide range of lesions and physiological conditions, including ACS and non-small vessels. ^3,4,5,6)^ However, large registries which provide information on the efficacy and safety of DCB for these currently expanded uses of DCB are scarce. The SeQuentPlease World Wide Registry reported in 2012 enrolled 2,095 patients. However, 77.2% of outcomes were related to in-stent restenosis, with only 572 lesions involved native coronary arteries.^13)^ Further, non-randomized studies have the drawback of being susceptible to confounding factors and bias. From this point of view, the present study provides significant advantages over previous registries. We enrolled 1,817 patients in a remarkably brief period. For example, recently reported EASTBOURNE registry of sirolimus DCB took approximately four years to complete. ^14)^ Longer enrollment periods can introduce selection bias as physicians’ case selection and techniques evolve over time. This confounding effect limits the interpretation of single-arm trials. By rapidly enrolling many patients, our study minimizes these biases, and is worth highlighting. The two types of DCB used are the only types approved in Japan, and comparative studies have confirmed that their performance does not differ. ^15)^ We therefore suggest that any influence of selection bias on DCB performance can be considered negligible.

### DCB therapy combined with an imaging modality

The 1-year TLF of 4.7% is consistent with the EASTBOURNE registry with a TLF for de novo lesions of 4.9%.^14)^ However, the present study included patients undergoing hemodialysis and those with complex lesions such as calcified lesions treated with atherectomy, reflecting patient outcomes in broader contemporary clinical practice.

Moreover, the key difference from previous studies is that the DCB procedure was performed using an imaging modality. Notably, the ULTIMATE III trial reported a benefit of imaging-guided DCB compared with angiography-guide DCB, but did not evaluate clinical endpoints as a surrogate. ^8)^ In this context, our present study may support and strengthen the findings of ULTIMATE III. A plausible benefit of imaging guided DCB is that it enables accurate assessment of vessel diameter and lesion length, allowing selection of the appropriate balloon size and reducing the risk of lesion mismatch with the DCB. Indeed, in ULTIMATE III, the imaging-guided group selected larger balloon and demonstrated higher angiographic indices.^7)^ Secondarily, imaging may allow the selection of a more appropriate preparation device, and accurate assessment of optimal lesion preparation than angiography. Indeed, so-called atherectomy was utilized as a preparation device in approximately 20% of the present cases. The risk of acute coronary artery occlusion in the early phase after balloon dilation has been identified as a major drawback, and coronary dissection after pre-dilatation has been considered an undesirable feature arguing against inadequate lesion preparation. However, recent studies based on imaging findings suggest that coronary artery dissection without blood flow impairment may lead to better outcomes with DCB.^16,17)^ Thus, the finding of coronary dissection as bad sign for coronary occlusion may be balanced by its being a good sign for positive remodeling, to some extent at least. In the EASTBOURNE registry and REC-CAGEFREE study conducted under angiographic guidance, 8.7% and 9.4% of patients, respectively, required rescue stenting due to inadequate outcomes after DCB. ^14,18)^ In contrast, in this study, although the DCB procedure was performed at the discretion of each physician rather than based on clear imaging criteria for optimal lesion preparation, rescue stenting was required in only 1.1% of cases. This finding may underscore the potential for imaging guidance to ensure safety and yield better outcomes with DCB.

### Risk factors associating with DCB outcomes

DM, hemodialysis patients, and ostial lesion were independently associated with higher TLF and cd-TLR. These factors are well-known risk factors for DES failure. It seems reasonable to consider that inadequate lesion preparation is a common mechanism for both DES and DCB failure. Furthermore, patients with DM or on hemodialysis may be at increased risk of accelerated intimal hyperplasia. On the other hand, hyperlipidemia and ACS were negative and positive risk factors for TLF, respectively, but were not risk factors for cd-TLR. This suggests that ischemic events may contribute to TLF in these subsets. It should be emphasized that ACS, calcified lesions, diffuse lesions, bifurcation lesions, and non-smaller vessels were not risk factors for cd-TLR. These findings are consistent with recent studies^3,5,6)^ and may suggest that DCBs should be actively considered in high-bleeding-risk patients for whom long-term DAPT is inappropriate.

However, DCB in the REC-CAGEFREEⅠ trial was inferior to DES in non-small vessels and interaction was observed. ^18)^ Similarly, to the REC-CAGEFREEⅠtrial, univariate analysis in our registry showed that non-small vessels were a risk factor for TLR, albeit that this difference was lost after adjustment for co-founders. The reason for these difference in results is unclear, but we speculate that several factors may be involved. The plaque burden is marked, the acute recoil will be large. The difference in acute lumen gains between DES and DCB may be significantly greater in non-small vessels due to vessel recoil. As a result, DES may provide greater benefit in large vessels. From this perspective, the benefits suggested by ULTIMATE III may be more pronounced in non-small vessels. The combined advantages of using an imaging modality—including selecting appropriate preparation devices and balloon sizes while confirming intraluminal dilatation—may be more pronounced in non-small vessels. In addition to these procedural differences, the differences in outcome may be also partly attributable to variations in lesion complexity and patient background. Finally, the DCB used in this study differed from that used in that study.

### Limitations

Several important limitations of this study should be mentioned. First, the study was conducted under a single-arm, open-label registry design, with all the limitations inherent to it. Therefore, caution is required in interpreting the findings of this study. Nevertheless, this concern was partly mitigated by the extremely short enrollment period, the all-comer approach, and the fact that all outcomes were determined by an independent clinical events committee based on prespecified criteria. Second, although outcomes of DCB were consistently good for all lesions, the present results were limited to cases in which lesion preparation was considered successful. In addition, the success or failure of lesion preparation depended on the judgement of the operator. Therefore, the results cannot be generalized to all patients scheduled for coronary intervention.

Generalization may require a specific definition of imaging guidance. In addition, confirmation requires a comparative study of imaging-guided DCB against DES. Third, the lack of core laboratories for angiography and imaging and the fact that qualitative and quantitative comparative analysis data are based on institutional reports may reduce objectivity in outcomes. The DCBs used were limited to paclitaxel-coated balloons and it is unclear whether similar results can be achieved with sirolimus-eluting balloons or other kind of paclitaxel DCB. Finally, the duration of DAPT was also left to the discretion of the physician, and although DCB is presumed to be effective in HBR cases, bleeding complications were not negligible in the present study. The appropriate duration of DAPT should be clarified.

## Conclusion

This large all-comer registry, which completed enrollment in a short period of time, demonstrated the efficacy and safety of paclitaxel DCB across various patients and lesions. DCB procedure conducted with the use of an imaging modality may enhance the safety and efficacy of DCB in daily practice.

## Acknowledgements

The authors thank Etsuyo Kitakata, and Akihiko Nishimura (Meditrix, Inc. Tokyo, Japan) for study management and Shiro Ueda (Medical Edge Co.) for statistical assistance.

Supplemental Material

Tables S1-S7.

References 1-6.

## Abbreviations and acronyms

ACS: Acute coronary syndrome
BARC: Bleeding academic research consortium
Cd TLR: Clinically driven target lesion revascularization
DAPT: Dual antiplatelet therapy
DCB: Drug-coated balloon
DES: Drug-eluting stent
DM: Diabetes mellitus
TLF: Target lesion failure

---

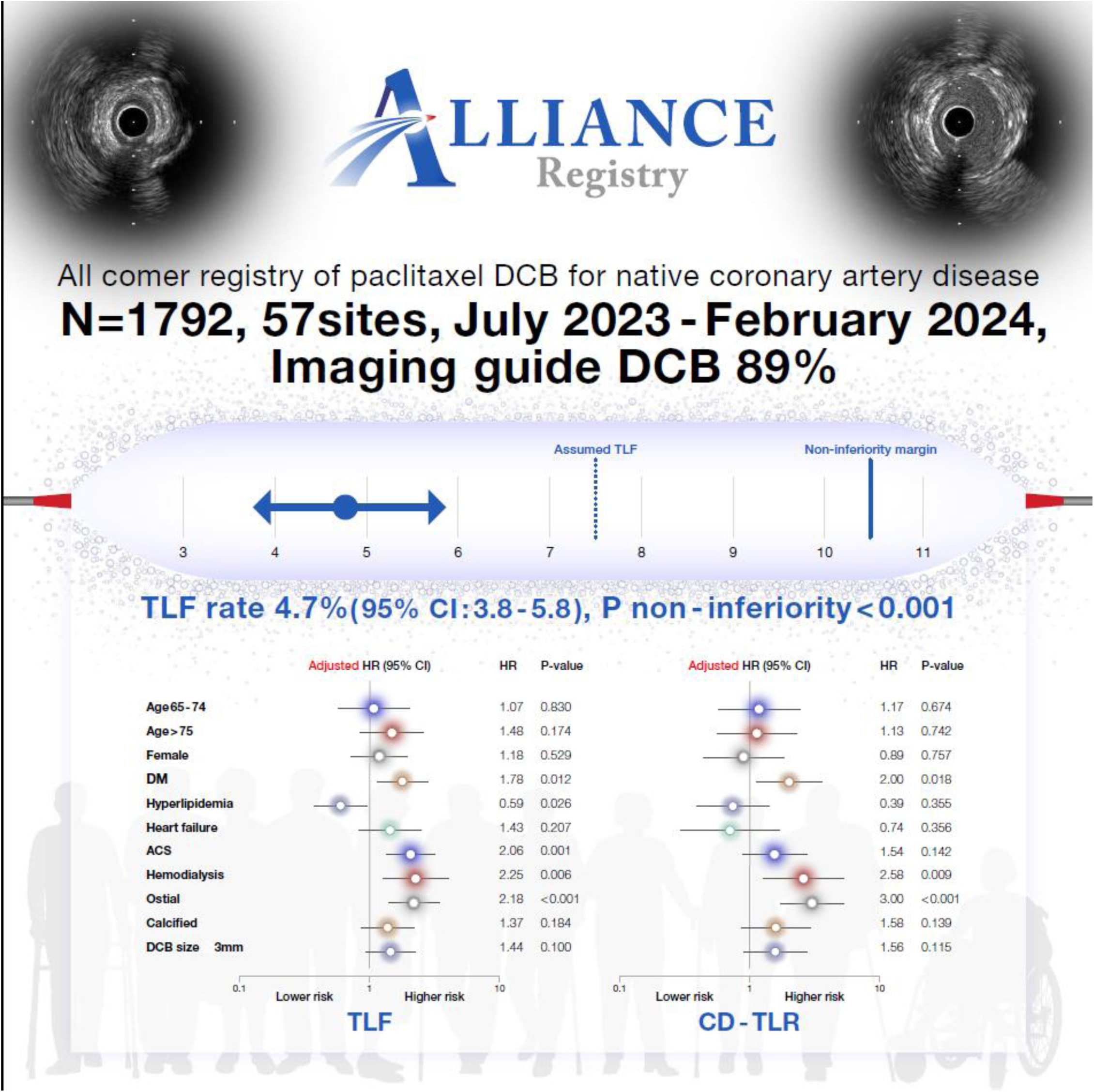
**Graphic abstract:** Registry overview and main findings.

